# A data driven epidemic model to analyse the lockdown effect and predict the course of COVID-19 progress in India

**DOI:** 10.1101/2020.06.01.20119388

**Authors:** B.K. Sahoo, B.K. Sapra

## Abstract

We propose a data driven epidemic model using the real data on the infection, recovery and death cases for the analysis of COVID-19 progression in India. The model assumes continuation of existing control measures such as lockdown and quarantines, the suspected and confirmed cases and does not consider the scenario of 2^nd^ surge of the epidemic due to any reason. The model is arrived after least square fitting of epidemic behaviour model based on theoretical formulation to the real data of cumulative infection cases reported between 24 March 2020 and 15 May 2020. The predictive capability of the model has been validated with real data of infection cases reported during May 15–30, 2020. A detailed analysis of model predictions in terms of future trend of COVID-19 progress individually in 18 states of India and India as a whole has been attempted. Infection rate in India as a whole is continuously decreasing with time and has reached 3 times lower than the initial infection rate after 6 weeks of lock down suggesting the effectiveness of the lockdown in containing the epidemic. Results suggest that India as a whole could see the peak and end of the epidemic in the month of July 2020 and January 2021. As per the current trend in the data, active infected cases in India may reach 2 lakhs at the peak time and total infected cases may reach around 14 lakhs. State-wise results have been discussed in the manuscript. However, the prediction may deviate particularly for longer dates, as assumptions of model cannot be met always in a real scenario. In view of this, a real time application (COV-IND Predictor) has been developed which automatically syncs the latest data from COVID19 dash board on daily basis and update the model input parameters and predictions of relevant results on daily basis. This application can serve as a practical tool for epidemic management decisions

## 1. Introduction

Corona virus disease 2019 (COVID-19) is an infectious disease caused by severe acute respiratory syndrome corona virus 2 (SARS-CoV-2) [1, 2], The disease was first identified in December 2019 in Wuhan, the capital of China’s Hubei province. Since then, the numbers of cases have spread to all over the world. On January 30, the World Health Organization [WHO] formally declared the outbreak of novel coronavirus as a Global Pandemic. As of June 01, 2020, a total of 6,152,160 cases are confirmed in more than 227 countries and 26 cruise ships. There are 3,142,964 active cases and 371,700 deaths [3].

The first case of the 2019–20 coronavirus pandemic in India was reported on January 30, 2020, originating from China. As of June 01, 2020, the Ministry of Health and Family Welfare has confirmed a total of 190,535 cases, 91,819 recoveries and 5,394 deaths in the country. India’s case fatality rate is relatively lower at 3.09%, against the global 6.63% as of 20 May 2020. Six cities account for around half of all reported cases in the country – Mumbai, Delhi, Ahmedabad, Chennai, Pune and Kolkata [4, 5]. On March 22, 2020, India observed a 14-hour voluntary public curfew followed by a nationwide lockdown since March 24, 2020, besides several other measures such as quarantine of the suspected cases, public health guidelines on social distancing, frequent hand washing and wearing face mask while stepping out of home for essential services.

Modelling and predicting the course of the outbreak in each region is important for the management and containment of the epidemic, and for balancing the impact from the public health vs. the economic crisis. Majority of COVID-19 epidemic models have originated from the SIR (Susceptible, Infected, and Recovered or Removed) model [6] and its many variations have been used in several countries, such as India [7], China [8, 9], Italy[10, 11] and Brazil [12], These SIR-type models are useful for policy-decision makers to know the potential impact of pandemic and for prompting them to take early actions to minimise the impact. However, subsequent to breakout of pandemic, more information is required for a detailed planning, such as peak arrival of the pandemic, the number of hospital beds needed at the peak time, and taking decision on relaxing/lifting the lockdown, and finally returning to normal living.

We propose a data driven epidemic model to predict the course of COVID-19 progress in India for the near term using the latest data on cumulative infection cases and removed cases due to recovery and death. The model has the advantage that it does not depend on the susceptible population, a key parameter required for SIR type models. However, it has the disadvantage that it cannot be used when the epidemic has just started and the data are limited.

## 2. Theoretical formulation and development of model

Let *N*(*t*) be the number of total infected cases at time *t*. The rate of change of total infected cases can be expressed as

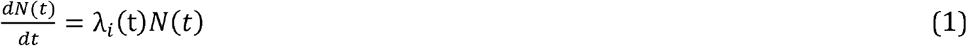

Where *λ_i_*(*t*) represents the infection rate at time *t*

Generally, infection rate represents the number of contacts per person per unit time and it decreases with various control measures such as quarantine, lockdown etc [13, 14]. Let us consider a scenario of continued lockdown till the epidemic comes to a near end. Also, it is assumed that the infection rate in the population is highest at start of lockdown which decreases exponentially with increase in lockdown period and finally approaches zero after a sufficient time.

With this, the transient variation of infection rate subsequent to lockdown can be written as

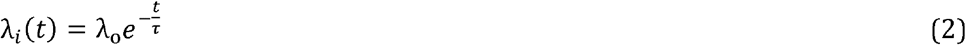

where λ*0* is the initial infection rate at time of implementing lockdown, τ is the characteristic time of decrease which depends upon the societal factor, the extent of implementation of the lockdown in the society, number of quarantine person, number of samples tested etc.

Substituting the expression for λ_*i*_(*t*), Eq (1) can be written as

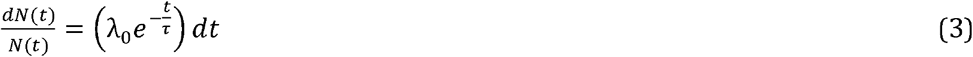

If *N*_0_ is the number of total infected cases at time of implementing lock down (*t* = *t_0_*], the solution to the above equation can be written as

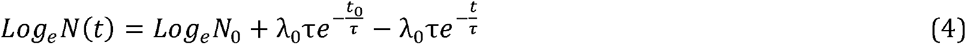

This can be re-expressed as

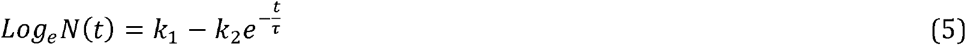

Where

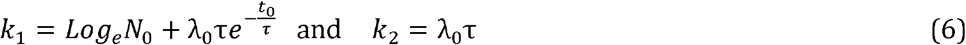

The number of infected cases, *N*(*t*)(using Eq 5 and 6) can be expressed as

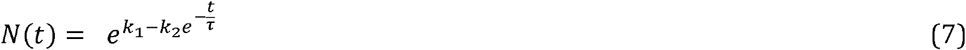

In real scenario, assumption of model cannot be met always. Hence there may a change in the trend of infections rate due to some reason (e.g. movement of migrant from one state to another), In such cases, there may be significant deviation between real infected cases and predicted infected cases. It is advisable to correct the deviation factor for the latest data (e.g. Moving average of deviation over a week) to the prediction model and re-estimate the projected data.

### 2.1. Daily new infected and removed cases

Differentiating Eq.(7) with respect to *t*, the number of *new* infected cases per day, *N_n_*(*t*) can be obtained as follows:

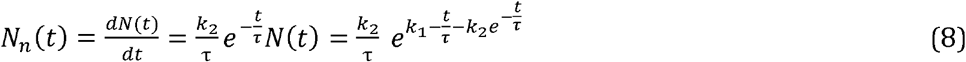

Since those who are admitted to thehospital either recover after a hospital stay of *T* days, or may dieafter a similar number of days, there should be a delayed relationship between number of new infected cases per day, *N_n_*(*t*) and number of removed cases per day, *N_r_*(*t*) due to recovery and death.

Hence, *N_r_*(*t*) can be related to *N_n_*(*t*) by the following relation

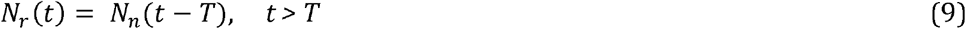

Where *T* is the mean recovery time of COVID-19 patients. This can be determined through time-lag correlation analysis between new cases and recovery cases where epidemic has nearly come to an end.

### 2.2. Active infected cases

Similarly, the number of infected active cases, *N_a_*(*t*) can be estimated by taking the difference between cumulative new infected cases and cumulative removed cases up to time *t* i.e.

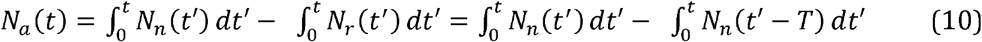

Simplifying, we get

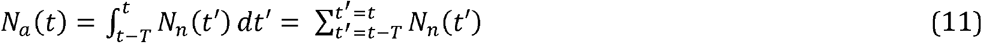

This indicates that the number of active infected cases at time *t*, is the sum of the number of daily new infected casesfor a period of *T* preceeding *t*.

### 2.3 Peak time of active infected cases

Maximum medical resources are required when the active cases attain maxima. Hence, predicting the maximum active cases and the time when this maximum will be attained is of utmost importance for planning and arrangement of medical resources such as number of hospital beds, ventilators, personal protective equipments for health care providers etc.

If *t_p_* is the time at which the active cases attaina peak, the occurrence of the peak is achieved when

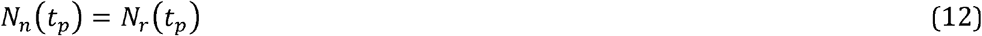

After attaining peak, the newly recovered and deceased cases start to exceed the newly infected cases. The demand for medical resources, such as hospital beds, isolation wards and respirators, starts to decrease beyond this peak.

Using equations (8) and (9), *N_r_*(*t*) can be expressed as

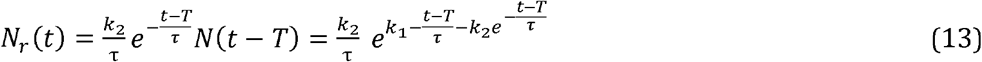

Taking the ratio of *N_n_*(*t*) and *N_r_*(*t*)as given in Eq.(8) and Eq.(13) respectively, and using condition given in Eq. (12) for the peak time, *t_p_* can be obtained as:

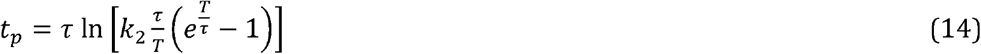

Eq.(14) can be used to obtain the time when active infected cases will attain peak with the information of characteristic time constant (τ), recovery time (*T*) and fitting parameter *k*_2_. Once the peak time *t*_p_ is estimated, the number of active infected cases at *t*= *t*_p_ can be estimated using Eq.(11).

## 3. Results and discussions

### 3.1. Estimation of model input parameters and validation

The first case of the 2019–20 corona virus pandemic in India was reported on January 30, 2020. India observed a nationwide lockdown since March 24, 2020 (55th day after 1^st^ case) to control the epidemic in addition to several other measures. As of June 01, 2020, the Ministry of Health and Family Welfare has confirmed a total of 190,535 cases, 91,819 recoveries and 5,394 deaths in the country. About 18 states have exceeded 100 confirmed cases as on May 01, 2020 [4, 5]. These states have been considered for model analysis and prediction. State-wise break up of confirmed cases as on March 24 (start date of lockdown) and as on June 01, 2020 and statistics of samples tested for these states is provided in Table 1.

**Table 1:** State-wise cumulative confirmed cases as on start date of lockdown and as on June 01, 2020, tested sample statistics and derived model parameters through least square fitting to time series data of respective states.

| State | Cumulative confirmed cases, $N(t)$ as on | | Samples tested per 1 million people | Confirmed cases out of 100 samples | Derived input parameters of Model (Eq.7)* | | | $R^2$ | Maximum % deviation from tested data (15-30 May) |
| --- | --- | --- | --- | --- | --- | --- | --- | --- | --- |
| | March 24 (lockdown start) | June 01, 2020 | | | $k_1$ | $k_2$ | Characteristic time constant, $\tau$ | | |
| All states (India) | 571 | 1,90,648 | 2799 | 5.1 | 14.1±0.1 | 19.9±0.5 | 53.9±1.9 | 0.999 | 8 |
| Maharashtra | 107 | 67655 | 3828 | 14.6 | 12.8±0.1 | 28.1±0.8 | 44.1±1.3 | 0.998 | 6 |
| Tamilnadu | 18 | 22333 | 6473 | 4.5 | 11.6±0.4 | 18.0±0.2 | 53.0±2.0 | 0.986 | 15 |
| Delhi | 30 | 19844 | 11821 | 9.3 | 11.7±0.3 | 18.6±1.0 | 53.9±4.2 | 0.989 | 10 |
| Gujarat | 34 | 16794 | 3215 | 8.2 | 10.1±0.0 | 108.7±7.1 | 22.2±0.5 | 0.993 | 9 |
| Rajasthan | 32 | 8831 | 5254 | 2.2 | 9.8±0.5 | 66.1±9.6 | 27.0±1.5 | 0.997 | 10 |
| Mandhya Pradesh | 7 | 8089 | 2046 | 4.8 | 8.8±0.1 | 115.3±12.7 | 19.2±0.8 | 0.982 | 10 |
| UttarPradesh | 35 | 8075 | 1266 | 2.8 | 9.2±0.2 | 43.7±3.8 | 27.2±1.4 | 0.997 | 4 |
| WestBengal | 9 | 5501 | 2079 | 2.7 | 9.3±0.3 | 35.0±3.1 | 34.0±2.5 | 0.995 | 5 |
| Bihar | 3 | 3807 | 636 | 5 | 11.1±1.1 | 40.1±0.3 | 46.4±11.5 | 0.983 | 20 |
| Andhra Pradesh | 8 | 3571 | 7033 | 1 | 8.7±0.4 | 27.9±7.8 | 31.2±6.0 | 0.981 | 10 |
| Karnataka | 41 | 3221 | 4448 | 1.1 | 11.5±1.2 | 26.3±7.2 | 53.1±4.1 | 0.989 | 12 |
| Telangana | 37 | 2698 | - |  | 11.1±0.0 | 35.0±26.9 | 52.0±0.3 | 0.986 | 7 |
| Jammu & Kashmir | 6 | 2446 | 12218 | 1.4 | 7.4±0.3 | 80.0±9.7 | 20.0±4.6 | 0.991 | 8 |
| Punjab | 29 | 2263 | 2928 | 2.6 | 8.1±0.5 | 370.0±18.3 | 16.0±6.0 | 0.980 | 11 |
| Haryana | 30 | 2091 | 4375 | 1.8 | 7.3±0.2 | 80.0±15.8 | 21.0±6.8 | 0.961 | 5 |
| Odisha | 2 | 1948 | 3381 | 1.3 | 9.1±1.2 | 60.0±10.5 | 34.1±18.0 | 0.978 | 18 |
| Assam | 0 | 1340 | 3117 | 1.2 | 11.1±1.1 | 70.0±13.0 | 45.1±3.7 | 0.994 | 20 |
| Jharkhand | 0 | 635 | 1781 | 1 | 8.9±0.2 | 40.2±19.1 | 45.1±1.3 | 0.973 | 10 |

\*Model input parameters should be re-estimated with latest data to make the predictions more accurate

The time series data of confirmed cases between March 24 and May 15, 2020 have been converted into logarithmic values as per requirement of the model (Eq.5) and least square fitted using Origin software. Fitted curve to the data of India as a whole is shown in Fig 1. Similar least square fitting exercise has been carried out for the selected states as well. The derived fitting parameters for the selected states and India as a whole are also presented in Table 1.

**Fig 1:**
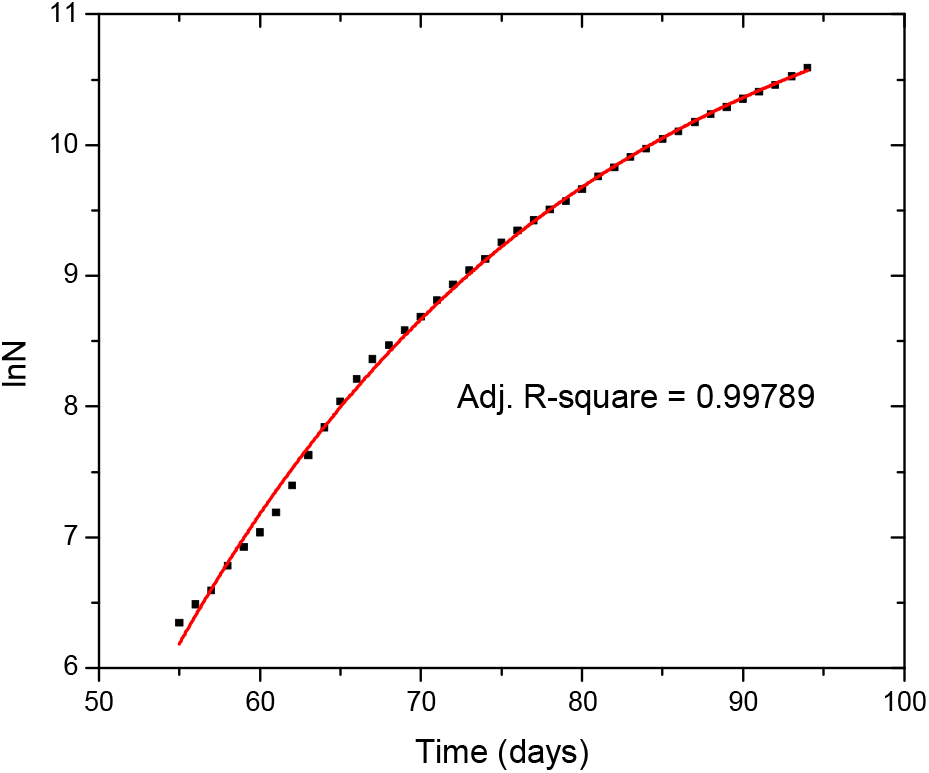
Least square fitting of model given in Eq. [7] to the data of cumulative infected cases [N] with time for India as a whole.

Subsequently, models for each state and India as a whole have been tested against the real data of confirmed cases reported during May 15–30, 2020 to find the deviation of predicted values and testing the validity of the model. The maximum percentage deviation of model predictions has been given in Table 1. The results shows that deviations are within 10 % in most of the states except a few states like Tamil Nadu, Bihar, Assam, Karnataka, Haryana and Odisha where it appears that a 2^nd^ surge is emerging. In these states, model prediction may not be accurate with the existing fitting parameters and needs to be updated using upcoming real data. It is important to note that the model is derived based on the certain assumptions as highlighted in model formulation section and does not consider scenario of 2^nd^ surge due to any reason. Hence, model parameters needs to be updated in such changed scenario.

### 3.2. Effect of lock down on infection rate

Using Eq.(1), one can express, the time-dependent infection rate as:

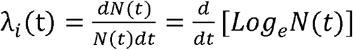

Using Eq.(2), the infection rate (per day) can be estimated by knowing the parameter λ_*0*_and τ. τ is the characteristic time obtained directly from least square fitting analysis and is given in Table 1. λ_*0*_ can be obtained as the ratio of fitting coefficient *k_2_* and τ.

Fig. 2 shows the plot of infection rate (predicted)and the real data of cumulative infected casesin India as a whole as a function of timet. Time, *t* = 0 represents the start date of lockdown (i.e 24 March 2020). As may be seen, there is a very good matching (< 10 % deviation) in the trend of predicted and real infection rate suggesting the validity of the model. Also, the falling infection rate suggests the effectiveness of the lock down in containing the epidemic. Initially the infection rate was around 0.15 per day which has come down to about 0.05 per day (about 3 times lower) after 6 weeks of lock down. If this trend continues, the predicted infection rate will reach one tenth of the initial infection rate (∼0.015) by about 12 weeks from start date of the lockdown. The characteristic time constant, τ, governs the decreasing trend of the infection rate. Higher this value, slower is the decrease in rate of infection. Table 1 provides the value of characteristic time constant for various states and results indicate that the time constant for most affected states of India such as Maharashtra, Tamil Nadu, Delhi and Karnataka is relatively higher in comparison to other states. Comparing the infection rate of India as a whole with countries such as USA, Italy, Germany and China, one can observe that the infection rate is quite low (∼ 2–3 times) in India. However, the decreasing trend is not as fast as USA, Italy, Germany and China [2], This may be due to low testing of samples at initial time in India.

**Fig 2:**
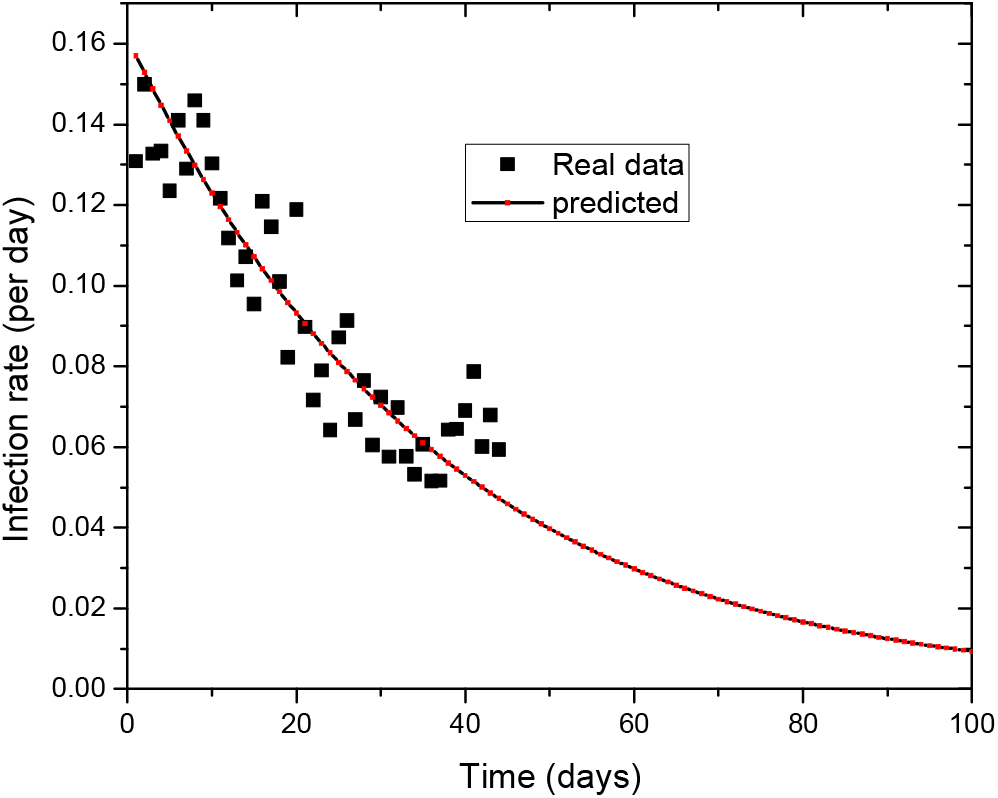
Trend of infection rate with days after lock down in India on 24 March 2020.

### 3.3. Time-lag correlation between new and removed cases and estimating mean recovery time

Now that the epidemic in Kerala appears to have come to an end, the data from this state has been used to perform cross correlation between daily new cases and removed cases due to recovery and death during the period March 14, 2020 and April 30, 2020. The plot of normalised correlation factor with respect to maximum value with different time lag is shown in Fig 3. As may be seen, the correlation is found to attain maximum when the time lag between them is 15 days i.e peak of new cases and removed cases is just lagged by 15 days. This is known as the mean recovery time, *T* for COVID patients. Typically for COVID-19 infection, the reported value of the mean recovery time varies from 14 to 16 days [2]. This recovery time of 15 days has been used for other states and India as a whole to estimate the peak of active infected cases.

**Fig 3:**
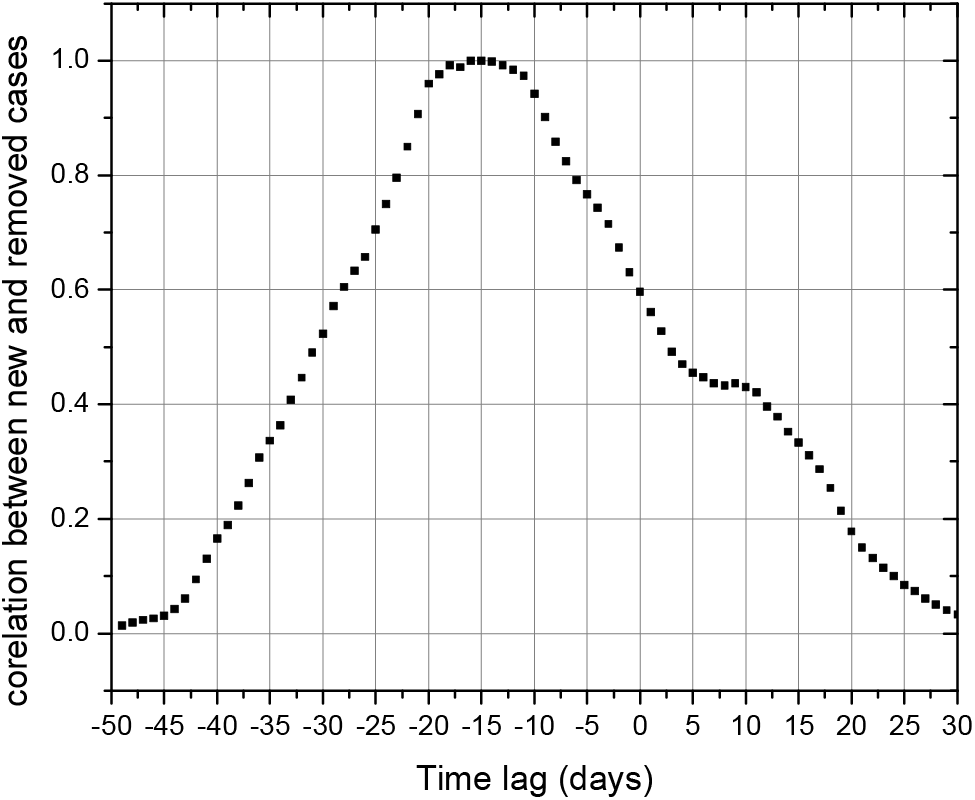
Time lag correlation analysis of daily new cases and removed cases due to recovery and death for the state of Kerala during the period March 14, 2020 and April 30,2020.

### 3.3. Prediction of daily, cumulative and active infected cases

Subsequent to estimation of mean recovery time and model parameters through least square fitting exercise, predictions have been made for daily, cumulative and active infected cases with time. In this analysis, January 30, 2020 is designated as time, *t* = 1 (date of 1^st^ reported case in India) and accordingly March 24, 2020 is t = 55 (start date of lock down). Predictions have been made only for *t*>55 till the cumulative infection cases attain saturation.

Fig 4 shows the plot of predicted mean, minimum, maximum of total infected cases, new daily cases and active infected cases in India with time. Fig 5, Fig 6 and Fig 7, show the plot of predicted total infected cases, new daily cases and active infected cases with time respectively in selected states of India. One can refer these plots to find the approximate time of peaking and near end of epidemic and number of active infected cases and saturation cases at peak and end time in various states of India and India as a whole.

**Fig 4:**
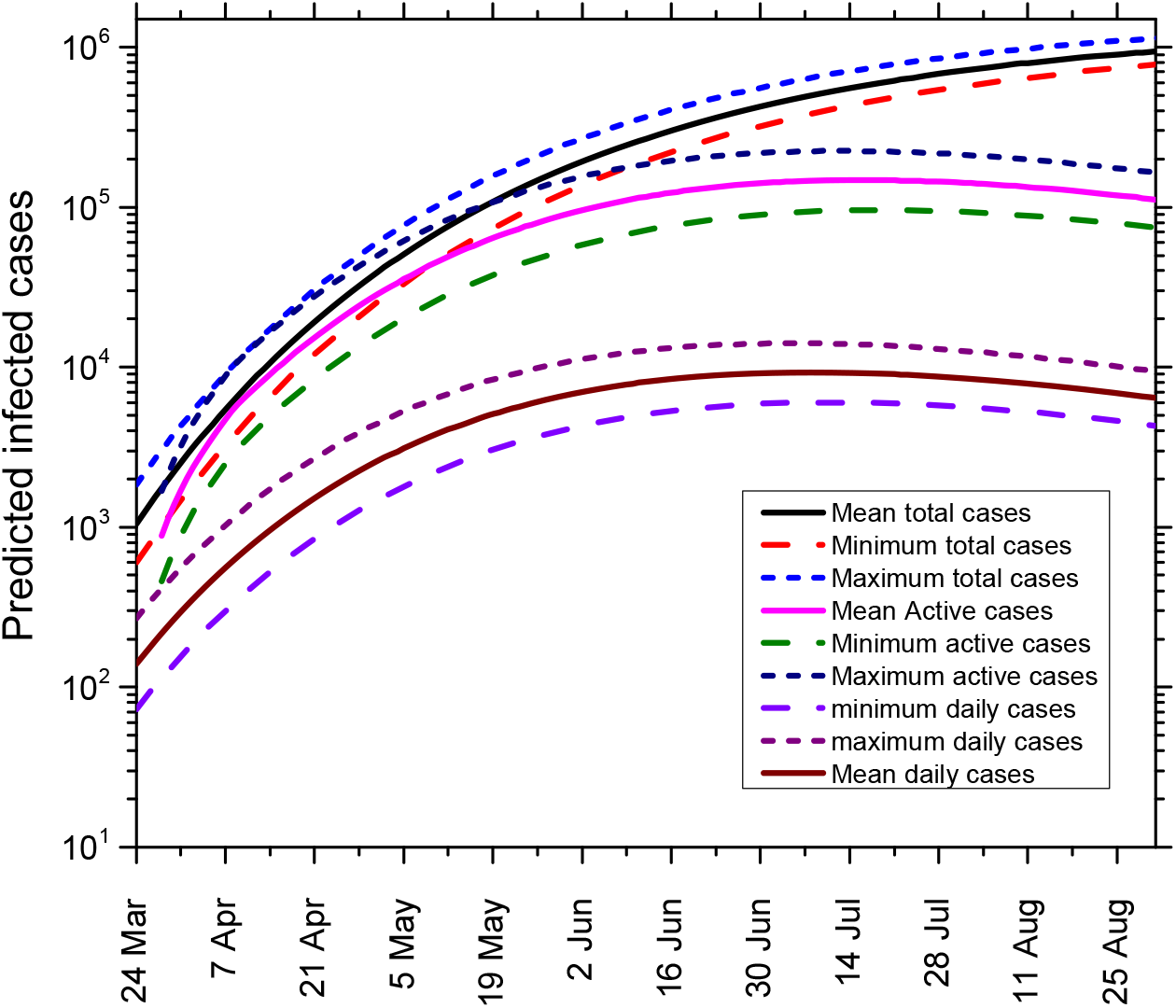
Predicted mean, minimum, maximum of total infected cases, new daily cases and active infected cases in India as a whole

**Fig 5.**
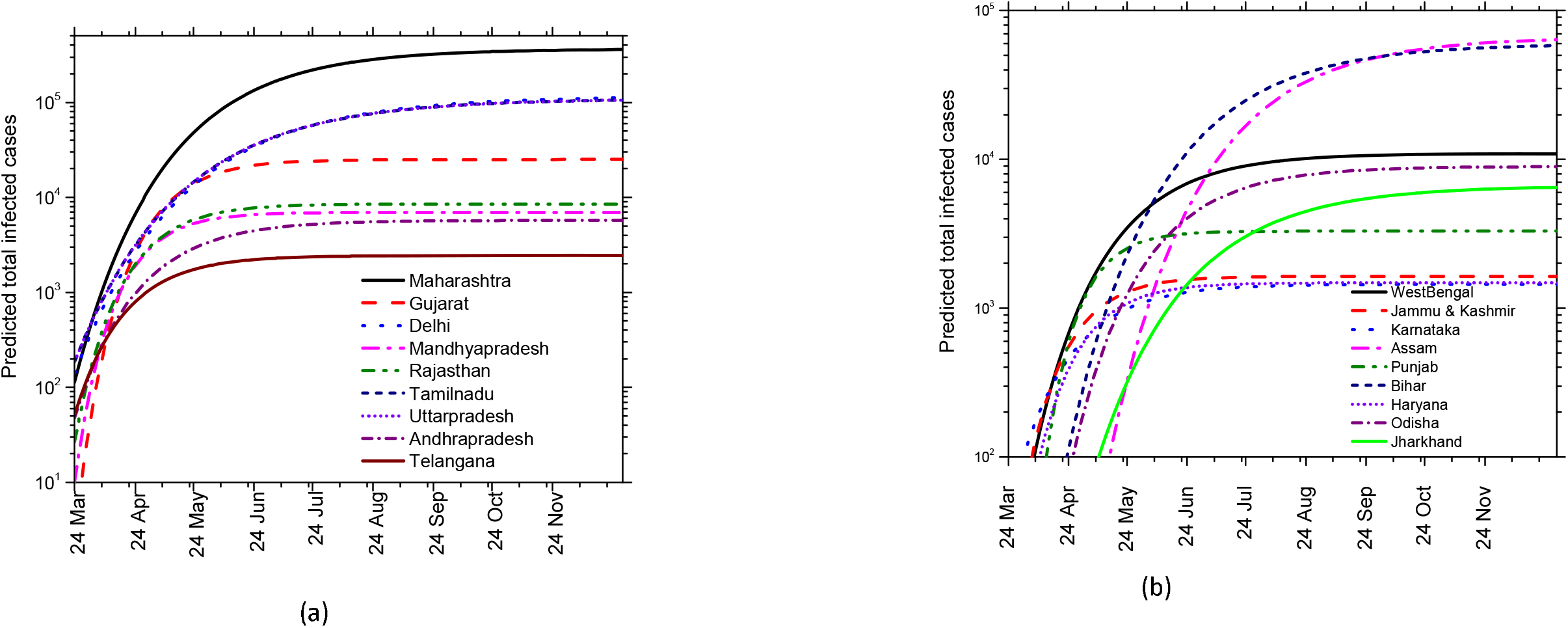
(a) and (b): Predicted total infected cases in various selected states of India

**Fig 6.**
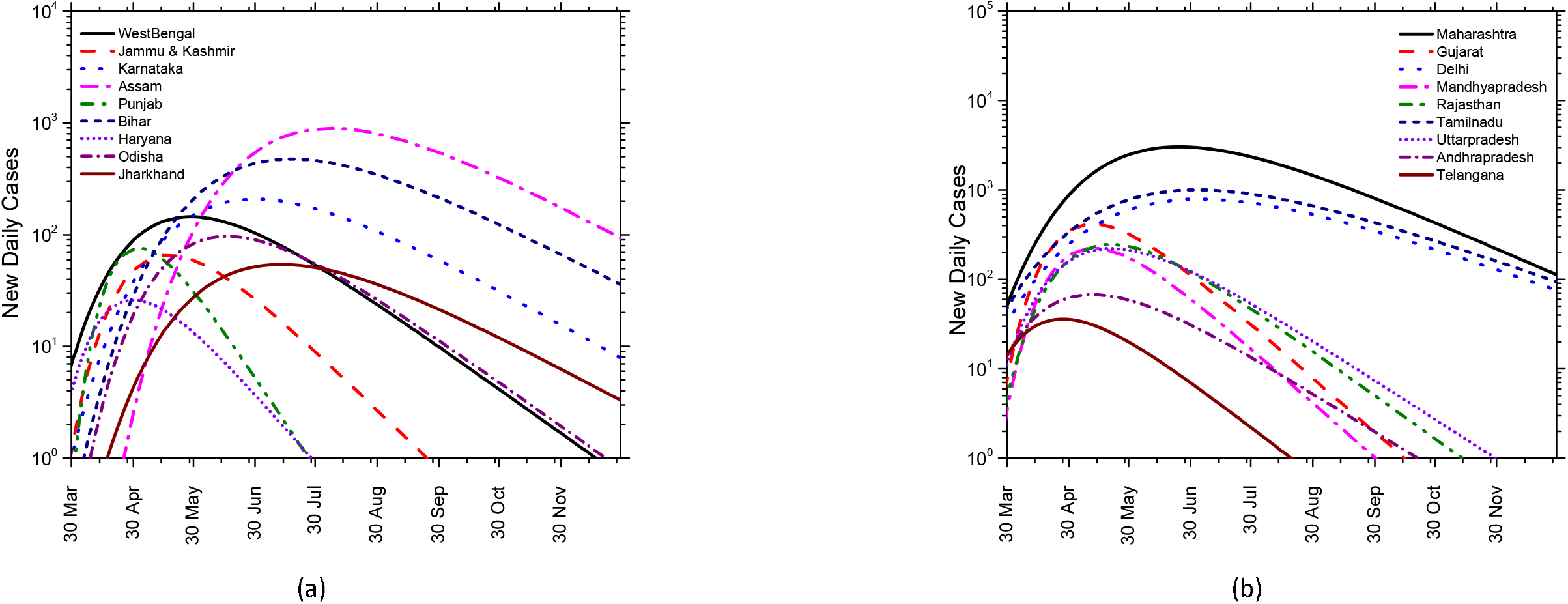
(a) and (b): Predicted total infected cases in various selected states of India

**Fig 7.**
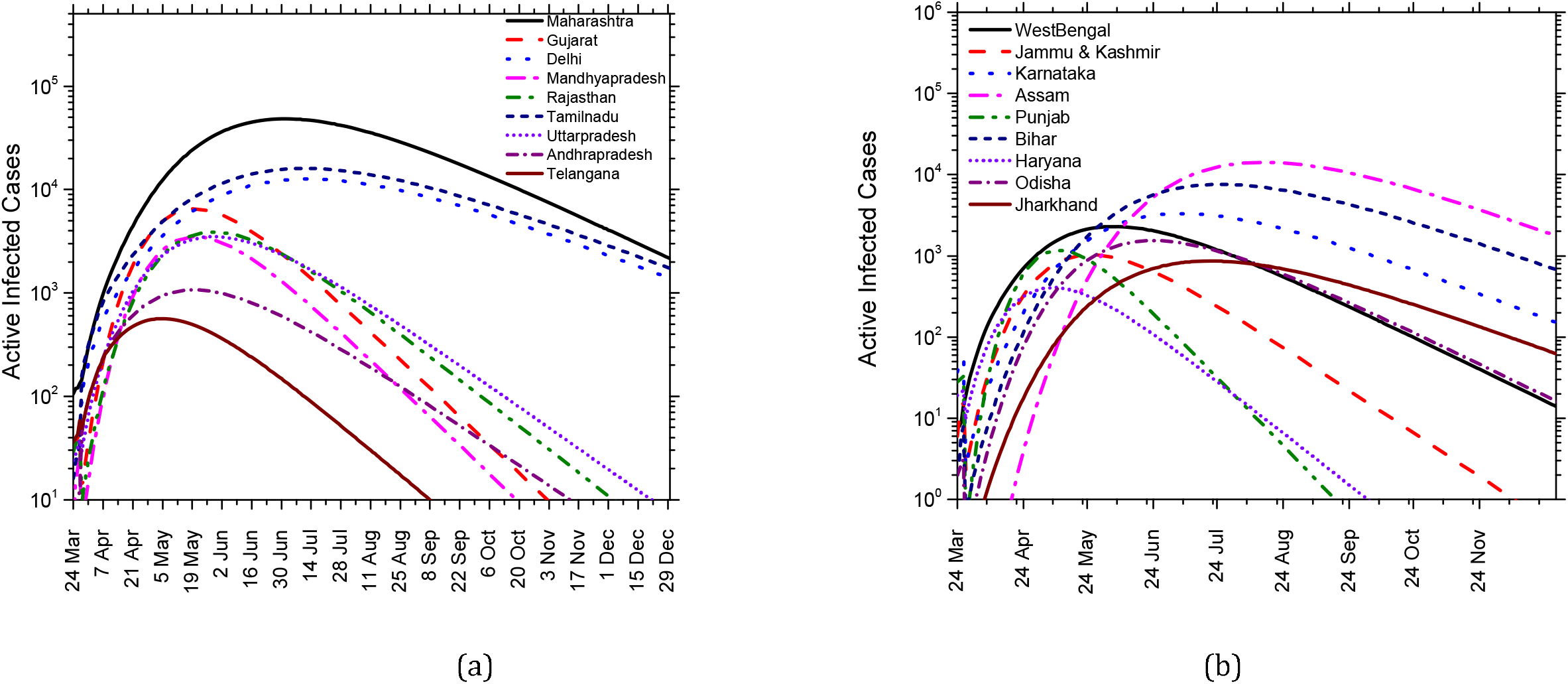
(a) and (b): Predicted active infected cases in various selected states of India

### 3.4. Prediction of peak and end time of epidemic and maximum active infected cases

Table 2 provides state-wise results of predicted time to reach peak of the epidemic, time to attain 99 % of the total infected cases (∼ end time of epidemic) and number of active and total saturation of infected cases with lower and upper bound value considering the error margin in the derived model input parameters. These results are very much useful for planning and arrangement of medical resources.

**Table 2:** State-wise predicted peak time, time to attain 99 % of the total expected cases and number of active and total saturation infected cases.

| State | Time to reach peak of the active infected case (MM-YYYY)* | Number of active infected cases at peak time* |  |  | Time to reach 99 % of total infected cases (MM-YYYY)* | Number of total expected infected cases* |  |  |
| --- | --- | --- | --- | --- | --- | --- | --- | --- |
|  |  | Most likely | Lower Bound | Upper Bound |  | Most likely | Lower Bound | Upper Bound |
| India as a whole | July 2020 | 1,70,437 | 1,48,543 | 1,98,235 | Jan 2021 | 13,10,305 | 11,86,453 | 14,25,385 |
| Maharashtra | July 2020 | 52,350 | 48,459 | 56,389 | Dec 2020 | 3,57,684 | 3,50,786 | 3,65,932 |
| Tamilnadu | July 2020 | 19,886 | 15,993 | 22,236 | Jan 2021 | 1,42,369 | 1,39,210 | 1,47,536 |
| Delhi | July 2020 | 16,832 | 13,457 | 20,854 | Jan 2021 | 1,38,346 | 1,20,906 | 1,58,657 |
| Karnataka | Aug 2020 | 15,378 | 10,718 | 20,157 | Jan 2021 | 1,04,216 | 93,622 | 1,15,213 |
| Assam | Sept 2020 | 14,236 | 10,357 | 18,359 | Jan 2021 | 1,15,439 | 1,05,423 | 1,25,547 |
| Bihar | Sept 2020 | 12,386 | 9,345 | 14,548 | Dec 2020 | 58,232 | 52,623 | 62,347 |
| Haryana | Sept 2020 | 11,567 | 6,678 | 18,523 | Oct 2020 | 54,567 | 46,987 | 67,975 |
| WestBengal | July 2020 | 5,638 | 4,378 | 7,471 | Oct 2020 | 52,065 | 44,849 | 58,578 |
| Gujarat | May 2020 | 6,790 | 5,897 | 7,229 | Aug 2020 | 35,145 | 30,345 | 39,127 |
| UttarPradesh | June 2020 | 5,188 | 3,697 | 6,358 | Oct 2020 | 34,740 | 28,459 | 40,702 |
| Rajasthan | June 2020 | 4,081 | 3,576 | 4,512 | Sept 2020 | 21,358 | 17,851 | 24,770 |
| MadhyaPradesh | June 2020 | 3,497 | 3,147 | 3,898 | Sept 2020 | 16,894 | 13,478 | 20,568 |
| Telangana | June 2020 | 2,739 | 2,512 | 2,912 | Dec 2020 | 19,564 | 17,214 | 21,121 |
| Jammu &Kashmir | July 2020 | 2,619 | 2,432 | 2,818 | Nov 2020 | 16,891 | 14,302 | 19,017 |
| Odisha | June 2020 | 1,536 | 1,434 | 1,678 | Nov 2020 | 8,851 | 7,812 | 10,112 |
| Jharkhand | June 2020 | 957 | 747 | 1,246 | Dec 2020 | 8,532 | 7,157 | 10,124 |
| AndhraPradesh | June 2020 | 1,514 | 1,110 | 1,820 | Oct 2020 | 5,731 | 4,657 | 7,964 |
| Punjab | May 2020 | 1,365 | 1,163 | 1,560 | Sept 2020 | 4,017 | 3,125 | 5,432 |

\* Results may deviate depending upon the change in the trend of the latest data. It is advisable to re-estimate above parameters in such scenario to make the predictions more accurate.

Results suggest that India as a whole could see the peak of the epidemic in the month of July 2020. Some of the states such as Kerala, Punjab, and Gujarat have already seen the peak of epidemic by this time and some of the states such as Uttar Pradesh, Madhya Pradesh, Rajasthan, Odisha, Jharkhand and Andhra Pradesh may see the peak in the month of June, 2020. The most affected states such as Maharashtra, Tamil Nadu, Delhi along with other states such as West Bengal and Jammu & Kashmir may see the peak of epidemic in the month of July, 2020. The remaining states Karnataka, Assam, Bihar, Haryana may see the peak in peak of the epidemic in later time (∼ September 2020) due to 2^nd^ surge of the epidemic in these states.

Results on active infected cases at peak time suggest that active infected cases for India as a whole may go up to 198 K (K stands for thousands, here onwards) with most likely number of cases around 170 K. The most affected states-Maharashtra and Tamil Nadu may see the active infected cases up to 56 K and 22 K respectively. Delhi, Karnataka, Assam, Bihar, Bihar could see the number of active infected cases in the range of 15K-20K at the peak time. Gujarat, West Bengal, Uttar Pradesh and Rajasthan may see the number of active infected cases between 5 K and 10 K at the peak time while remaining states may see the number of active infected cases below 5 K.

Time to reach 99 % of the total expected infected cases is considered as the end of the epidemic by which most of the active infected cases have recovered. Results suggest that India as a whole could see the end of the epidemic in the month of Jan, 2021 with a total cases reaching upto 14 lakhs. Gujarat could see end of the epidemic in the month of August 2020. Some of the states such as Madhya Pradesh, Rajasthan, Punjab could see the end of the epidemic by September 2020 and States such as West Bengal, Haryana Uttar Pradesh and Andhra Pradesh could see the end of the epidemic in the month of October 2020. Odisha and Jammu&Kashmir could see the end of epidemic in the month of November 2020 while Maharashtra, Jharkhand, Bihar and Telangana could see the end of epidemic in December 2020. The remaining states could see the end of epidemic in January 2021.

As it is well known, model assumptions cannot be met always in real scenarios and hence the prediction may deviate depending upon change in the trend in the data due to several factors such movement of migrant workers, epidemic control measures and its extent of implementation by each states and national governments. In this context, a real time application (COV-IND Predictor) has been developed by implementing the above model in a Google sheet which automatically syncs the latest data from COVID19 dash board [5] on daily basis and update the model input parameters and predictions of relevant results on daily basis. The application can be accessed from the link: https://docs.google.com/spreadsheets/d/1fCwgnQdz4J0YWVDHUcbEW1423wOJjdEXm8TqJDWNAk/edit?usp=sharing

It is advisable to check the latest predictions from the above link which gives a more reliable prediction of the COVID-19 scenario in India as a whole and in individual selected state based on the latest trend in the COVID-19 infected cases.

## 4. Conclusion

We propose a data-driven model to track and predict the course of the epidemic. Many parameters characterizing an epidemic can be determined from the model using the available latest data which can be validated by a few real data sets. Subsequently, the model can be used for predictions. This presented approach could be applied not just to the current Covid-19 epidemic, but also, in general, to future epidemics. The model gives best predictions with online type predictor, utilising latest data to update the model input parameters periodically and predict the course of epidemic for next two weeks. The model is of special significance for predicting the approximate peak time and end time of the epidemic so as to keep a readiness for maximum resources during the peak time. The model is able to well capture the observed decrease in the infection rate post lockdown, thus confirming the effectiveness of lockdown in containing the epidemic. The model has been implemented in a Google sheet which can serve as a practical tool for epidemic management decisions such as relaxing lockdown, quarantine institutions and medical resource planning and balancing the impact from the public health vs. the economic crisis [15–18].

## Data Availability

All real data on COVID -19 upto June 01 have been retrieved from https://www.covid19india.org/

## Acknowledgement

Authors would like to thank Shri Suresh Babu RM, Associate Director, Health, Safety and Environment Group of BARC for his valuable suggestions and inputs during internal review of the manuscript

